# Undeclared Kratom Alkaloids and Tianeptine in Products Labeled as Cat’s Claw

**DOI:** 10.64898/2026.10.16.26365420

**Authors:** Bharathi Avula, Kumar Katragunta, Kiran Kumar Tatapudi, Ji-Yeong Bae, Amar G. Chittiboyina, Ikhlas A. Khan

## Abstract

**Background:** High-concentration 7-hydroxymitragynine (7-OH) products have been described by the US Food and Drug Administration as an emerging public health concern, and in August 2026 the Drug Enforcement Administration temporarily placed mitragynine pseudoindoxyl (MPI), MGM-15 (dihydro-7-hydroxymitragynine), and MGM-16 in schedule I. Since July, Utah and Tennessee officials have reported overdoses, deaths, and laboratory findings involving tablets labeled as cat’s claw (*Uncaria tomentosa*), an herbal supplement without known opioid activity. Chemical data on such products are scarce, and labels do not tell consumers what the products actually contain, including semi-synthetic opioids.

**Methods:** In a cross-sectional analysis, we purchased 46 chewable or unit-dose products labeled as containing cat’s claw from online retailers and three different gas stations in Mississippi during September 2026. The products represented 17 brands. Methanol extracts were analyzed by ultra-high-performance liquid chromatography with photodiode array detection and compared with a methanol extract of botanically verified *U tomentosa* bark. Liquid chromatography quadrupole time of flight mass spectrometry identified additional compounds, and compounds were quantified against authentic reference standards. Proportions are reported with exact (Clopper-Pearson) 95% CIs.

**Results:** None of the 46 products declared kratom alkaloids or their derivatives, yet all contained at least one component. Mitragynine was quantified in 41 products (89%; 95% CI, 76% to 96%), 7-OH in 35 (76%; 95% CI, 61% to 87%), MGM-15 in 24 (52%; 95% CI, 37% to 67%), and MPI in 22 (48%; 95% CI, 33% to 63%). A total of 39 products (85%; 95% CI, 71% to 94%) contained MPI or MGM-15, which are temporarily controlled in schedule I. Tianeptine, an antidepressant not approved in the US, was present in five products (11%; 95% CI, 4% to 24%) at 27.9 to 40.2 mg per unit. The 7-OH content ranged from 0.01 to 95.4 mg per unit. Eight products carrying claims such as “NO KRATOM and No 7-OH” or “NO 7-OH” contained mitragynine and 7-OH.

**Conclusions:** Labels gave consumers no reliable guide to exposure. Clinicians evaluating unexplained opioid toxicity after use of a cat’s claw product should consider kratom-derived opioids and tianeptine. Regulators may need product testing and label enforcement in addition to scheduling of individual compounds.

## Introduction

US Food and Drug Administration has described high-concentration 7-hydroxymitragynine (7-OH) products as an emerging public health concern.^1,2^ In August 2026, the Drug Enforcement Administration temporarily placed mitragynine pseudoindoxyl (MPI), MGM-15 (dihydro-7-hydroxymitragynine), and MGM-16 (a fluoro analog of MGM-15) in schedule I.^3^ Since July, Utah and Tennessee officials have reported overdoses, deaths, and laboratory findings involving tablets labeled as cat’s claw (*Uncaria tomentosa*), an herbal supplement without known opioid activity.^4,5^ Chemical data on such products are scarce. We measured the composition of products labeled as cat’s claw and compared it with their labels.

## Methods

In a cross-sectional analysis, we purchased 46 chewable or unit-dose products labeled as containing cat’s claw from online retailers and three different gas stations in Mississippi during September 2026 (Supplement Information, **Table S1**). The products represented 17 brands, including multiple flavors per brand. A few products were specifically labeled and marketed as “NO KRATOM” or “NO 7-OH.”

Methanol extracts of these products were quantified by ultra-high-performance liquid chromatography (UHPLC) with photodiode array detection (PDA), and their profiles were compared with a methanol extract of botanically verified *U. tomentosa* bark [12162] to assess the presence oxindole alkaloids. Liquid chromatography quadrupole time of flight mass spectrometry (LC-QToF-MS) identified additional compounds by retention time, accurate mass, and fragment ions, (LOD, 1 to 100 ng/mL). Nine compounds (mitragynine, 7-OH, MPI, 3-dehydromitragynine, MGM-15, yohimbine, tianeptine, melatonin and caffeine) were quantified against authentic reference standards^6^ and reported as milligrams per unit. Proportions are reported with exact (Clopper-Pearson) 95% CIs. The study analyzed commercial products and involved no human participants. We followed the STROBE reporting guideline for cross-sectional studies.

## Results

None of the 46 cat’s claw products declared kratom alkaloids or their derivatives, yet all contained at least one component (Table). Mitragynine was quantified in 41 products (89%; 95% CI, 76% to 96%), 7-OH in 35 (76%; 95% CI, 61% to 87%), MGM-15 in 24 (52%; 95% CI, 37 to 67%), and MPI in 22 (48%; 95% CI, 33% to 63%). A total of 39 products (85%; 95% CI, 71% to 94%) contained MPI or MGM-15, which are temporarily controlled in schedule I. Tianeptine, an antidepressant not approved in the US,^7^ was present in five products (11%; 95% CI, 4% to 24%) at 27.9 to 40.2 mg per unit. One product also contained yohimbine, melatonin, and lidocaine. Although it was not quantified, rhynchophylline was detected in 13 products (28%; 95% CI, 16% to 43%), but none showed the exclusive oxindole alkaloid profile of authentic *U. tomentosa* bark.

**Table.** Compounds Detected in 46 Products Labeled as Cat’s Claw.

| Compound | Products, No. (%)<br>(N = 46) | Concentration, median<br>(range), mg per unit<sup>b</sup> |
| --- | --- | --- |
| Mitragynine | 41 (89) | 0.8 (0.02 to 32.5) |
| 7-Hydroxymitragynine | 35 (76) | 0.7 (0.01 to 95.4) |
| MGM-15<sup>a</sup> | 24 (52) | 21.6 (3.1 to 28.7) |
| Mitragynine pseudoindoxyl (MPI)<sup>a</sup> | 22 (48) | 0.7 (0.03 to 1.7) |
| Either MGM-15 or MPI<sup>a</sup> | 39 (85) | NA |
| 3-Dehydromitragynine | 46 (100) | 0.15 (0.001 to 2.5) |
| Tianeptine | 5 (11) | 39.3 (27.9 to 40.2) |
| Caffeine | 14 (30) | 2.6 (0.1 to 3.5) |
| Rhynchophylline | 13 (28) | NQ |

Abbreviation: NA, not applicable; NQ, not quantified
<sup>a</sup>Temporarily controlled in schedule I (August 2026); <sup>b</sup>Summarized among products in which the compound was quantified.

Composition varied widely (Supplement Information, **Table S2**). The 7-OH content ranged from 0.01 to 95.4 mg per unit; 16 products contained 22.2 mg or more, and 12 contained 40 mg or more. MGM-15 ranged from 3.1 to 28.7 mg per unit and MP from 0.03 to 1.7 mg per unit. Three products contained mitragynine at 31.2 to 32.5 mg per unit. Within a single 5-flavor product line, 7-OH content ranged from 45.2 to 84.2 mg per unit. Eight products carrying claims such as “NO KRATOM and No 7-OH” or “NO 7-OH” contained mitragynine, 7-OH, and 3-dehydromitragynine. Labeled ingredients were not detected in 41 products.

## Discussion

All 46 products labeled as cat’s claw contained undeclared kratom-derived alkaloids, 85% contained schedule I semisynthetic opioids, and five contained the antidepressant tianeptine (Supplemental Information, **Figure S1**). Concentrations of 7-OH reached 95.4 mg per unit, composition differed among flavors of the same brand, and “no kratom” claims did not guarantee the absence of kratom alkaloids. Labels therefore gave consumers no reliable guide to exposure. Because tianeptine, MPI, and MGM-15 act as mu opioid receptor agonists,^3,8^ the findings are consistent with the opioid toxicity that Utah and Tennessee officials have attributed to similarly branded products.^4,5^ Poison center exposures to tianeptine have also increased.^9^

Ingredient identity, not the plant name on the label, determines the authenticity and its biological effects. The eight products labeled as free from kratom or free from 7-OH also contained kratom-derived alkaloids, including 7-hydroxymitragynine at concentrations far above those of the leaf,^6^ along with other semi-synthetic opiates. Only five of the 46 products contained *U. tomentosa* alkaloids, and each also possessed kratom alkaloids or their semi-synthetic derivatives, suggesting deliberate addition rather than botanical origin (Supplemental Information, **Table S3**). Clinicians evaluating unexplained opioid toxicity after use of a cat’s claw product should consider kratom-derived opioids and tianeptine. Regulators may need product testing and label enforcement in addition to scheduling of individual compounds.

Limitations include 46 products from 17 brands at one time point, no clinical outcome data, unassessed batch variability, and a targeted analyte panel that excludes MGM-16. We did not link any product to a specific case.

## Data Availability

All data produced in the present study are available upon reasonable request to the authors

## Declarations

### Funding/Support

Center for Food Safety and Applied Nutrition, US Food and Drug Administration, grant 5U01FD004246 (Science Based Authentication of Botanical Ingredients).

### Role of the Funder/Sponsor

The US Food and Drug Administration supports the National Center for Natural Products Research (NCNPR) through long standing cooperative agreement to ensure the quality and safety of botanicals in dietary supplements.

### Conflict of Interest Disclosure

None

### Ethics Statement

The study analyzed commercial products and involved no human participants.

### Data Availability

The methods for quantification, reference standards, per-product quantitative data, and mass fragmentation data are provided in the Supporting Information.

### Author Contributions

**BA:** methodology, validation, formal analysis, data curation, writing of the original draft, and review and editing; KK: product purchase, sampling, validation, quantitative measurements, and data curation; KKT: method development, qualitative and quantitative measurements; J-YB: method development, qualitative and quantitative measurements; CAG: Data analysis, statistics, writing of the original draft, review and editing and funding acquisition; IAK: supervision, project administration, and funding acquisition.

## Notes

### Competing Interest Statement

The authors have declared no competing interest.

## References

1. US Food and Drug Administration. Hiding in plain sight: 7-OH products. Updated July 13, 2026. https://www.fda.gov/news-events/public-health-focus/hiding-plain-sight-7-oh-products. Accessed October 7, 2026.

2. Gour A, Kanumuri SRR, Mukhopadhyay S, et al. Mislabeling and status of semisynthetic kratom-derived products in the US market. Drug Test Anal. 2026;18:1286-1295. doi:10.1002/dta.70122.

3. US Drug Enforcement Administration. Schedules of controlled substances: temporary placement of mitragynine pseudoindoxyl, MGM-15, and MGM-16 in schedule I. Fed Regist. August 26, 2026. https://www.federalregister.gov/documents/2026/08/26/2026-17429/schedules-of-controlled-substances-temporary-placement-of-mitragynine-pseudoindoxyl-MGM-15-and. Accessed October 7, 2026.

4. Utah Department of Health and Human Services. Utah health officials warn of fatal overdoses involving illegal “cat’s claw” and kratom product. Published October 5, 2026. https://dhhs.utah.gov/featured-news/utah-health-officials-warn-of-fatal-overdoses-involving-illegal-cats-claw-and-kratom-product/. Accessed October 7, 2026.

5. What is “cat’s claw” and why is Tennessee law enforcement warning public about it containing illegal compound? WSMV. September 14, 2026. https://www.wsmv.com/2026/09/14/what-is-cats-claw-why-is-tennessee-law-enforcement-warning-public-about-it-containing-illegal-compound/. Accessed October 7, 2026.

6. Avula B, Ali SK, Tatapudi KK, et al. Quantitative analysis of 7-hydroxymitragynine in commercial kratom products and its stability under chemical and physiological conditions. Phytochem. 2026;247:114871. doi:10.1016/j.phytochem.2026.114871

7. US Food and Drug Administration. Tianeptine in dietary supplements. https://www.fda.gov/consumers/health-fraud-scams/tianeptine. Accessed October 7, 2026.

8. Gassaway MM, Rives ML, Kruegel AC, et al. The atypical antidepressant and neurorestorative agent tianeptine is a mu-opioid receptor agonist. Transl Psychiatry. 2014;4(7):e411. doi:10.1038/tp.2014.30

9. Quadir M, Rine NI, Badeti J, et al. Tianeptine exposures reported to United States poison centers, 2015-2023. J Med Toxicol. 2025;21(1):30–41. doi:10.1007/s13181-024-01053-6

