## Supplemental data for "Undeclared Kratom Alkaloids and Tianeptine in Products Labeled as Cat’s Claw"

**Table of Contents:**

1. Methods for quantification of six alkaloids and tianeptine 02
2. Preparation of reference standards and calibration solutions and product samples 02
3. Instrumental conditions 03
4. Validation 03
5. References 04
6. **Table S1**. Lot or batch numbers, and other identifying information for the studied products 05
7. **Table S2**. Quantitative results of target alkaloids 14
8. **Figure S1**. Composition of measured compounds 16
9. **Table S3**. Mass spectral features of detected compounds in 46 products and cat's claw 17

**Methods for Quantification of Six Alkaloids and Tianeptine**

### 1. Chemicals and reagents

HPLC- grade acetonitrile, methanol, and LC-MS-grade formic acid were purchased from Fisher Scientific (Fair Lawn, NJ). Water was obtained from a Milli-Q-Gradient system (Millipore, Billerica, MA).

### 2. Preparation of reference standards and samples

#### 2.1 Preparation of reference standards, sources and calibration solutions

Mitragynine, paynantheine, speciogynine, and speciociliatine were isolated from botanically verified *Mitragyna speciosa* material at the National Center for Natural Products Research (NCNPR), University of Mississippi. Mitragynine served as the starting material for the synthesis of 7-hydroxymitragynine (7-OH) [1], 3-dehydromitragynine, and dihydro-7-hydroxymitragynine (MGM-15) [2] by previously reported methods. The identities of the isolated and synthesized compounds were confirmed by nuclear magnetic resonance (NMR) spectroscopy and high-resolution mass spectrometry (HRMS), and the spectroscopic and mass spectral data were consistent with previously reported literature values [3].

Mitragynine pseudoindoxyl, rhynchophylline, and tianeptine were purchased from Cayman Chemical (Ann Arbor, MI). Caffeine, theanine, melatonin, yohimbine, corynantheine, and corynantheidine were purchased from Sigma-Aldrich (St. Louis, MO). Lidocaine was purchased from MP Biomedicals (Solon, OH). All reference standards had stated purity above 96%.

Individual stock solutions of the seven quantitative reference standards (mitragynine, 7-OH, 3-dehydromitragynine, mitragynine pseudoindoxyl, MGM-15, tianeptine, and caffeine) were prepared in methanol at 2 mg/mL. Five-point calibration curves were prepared in methanol over the concentration range of 0.5 to 50 µg/mL. The remaining reference compounds were used for qualitative analysis and confirmation of compound identity and were not included in the quantitative calibration curves.

Authenticated *Uncaria tomentosa* bark reference material, used to establish the alkaloid profile of authentic cat's claw, was obtained from Missouri Botanical Garden (12162) and confirmed by the taxonomist.

#### 2.2 Preparation of working solutions of commercial cat's claw products in tablet form

A total of 46 products representing 17 brands, including different flavors or formulations within a brand, were analyzed. Each product was assigned a unique NCNPR repository code (#5443PR, 5445PR, 5469PR–5483PR, 5487PR–5515PR). Before analysis, the authors documented the product name, lot number, recommended serving size and instructions, Supplement Facts and other ingredient statements, manufacturer or distributor information, purchase source, and label claims.

All products were in tablet form. Tablets were weighed, finely ground, and homogenized with a mortar and pestle. Approximately 50 mg of each homogenized sample and a quantity equivalent to the average tablet weight were weighed separately into individual centrifuge tubes. Each sample was then extracted separately with 2.0 mL of methanol. The mixture was sonicated for 30 minutes and centrifuged for 15 minutes at 959 × *g*. The clear supernatant was transferred to a 10 mL volumetric flask. The extraction was repeated four times, and the supernatants were combined. The final volume was adjusted to 10 mL with methanol and mixed thoroughly. All extracts were filtered through a 0.45 µm polytetrafluoroethylene (PTFE) membrane filter before analysis.

### 3. Instrumental conditions

#### 3.1 Liquid chromatography-quadrupole time-of-flight mass spectrometry (LC-QToF-MS) for qualitative analysis

Qualitative analysis followed the previously reported methods [4-5] on an Agilent 1290 UHPLC coupled to a QToF-MS with ESI for compound identification and confirmation by accurate mass and diagnostic ions. Separation was achieved on a Poroshell 120 EC-$C_{18}$ column (2.1 × 150 mm, 2.7 µm) with 0.1% formic acid in water and acetonitrile at 0.2 mL/min. Mass spectra were acquired in positive-ion mode over *m/z* 50 to 1300, with accurate-mass calibration using reference ions at *m/z* 121.0509 and 922.0098. Compound identities were confirmed by comparison with authentic reference standards and their characteristic MS/MS fragmentation patterns. Limits of detection ranged from 1 to 100 ng/mL.

#### 3.2 Ultra-high-performance liquid chromatography with photodiode array detection (UHPLC-PDA) for quantitative analysis

Quantitative analysis of 7-OH, mitragynine, and mitragynine pseudoindoxyl used the validated LC–PDA method reported by Avula et al. (2026) [3]. Samples were extracted with methanol, filtered, and analyzed by chromatographic separation coupled with PDA detection, and these compounds were quantified as described earlier [3]. With the same analytical approach, four additional compounds (3-dehydromitragynine, MGM-15, tianeptine, and caffeine) were validated and then quantified in the samples. Based on the expected analyte concentrations in the products, sample extracts were diluted 10-fold to 1000-fold before analysis so that measured concentrations fell within the validated calibration range.

### 4. Validation procedure

The UHPLC-PDA method was validated for selectivity, specificity, linearity, sensitivity, limit of detection (LOD), limit of quantification (LOQ), accuracy, precision, and solution stability in accordance with the applicable International Council for Harmonisation (ICH) guidelines [6].

The LOD and LOQ were determined by serial dilution of standard solutions of each analyte and were defined as the lowest concentrations giving signal-to-noise (S/N) ratios of approximately 2 to 3 and 10, respectively. A five-point calibration curve was prepared for 3-dehydromitragynine, MGM-15, tianeptine, and caffeine. The curves were linear over the concentration range evaluated, with correlation coefficients ($r^2$) >0.99. The limits of detection (LODs) and limits of quantification (LOQs) for 3-dehydromitragynine and MGM-15 were 100 and 250 ng/mL, respectively, and those for caffeine were 25 and 100 ng/mL, respectively. For tianeptine, the LOD and LOQ were 250 and 500 ng/mL, respectively. All standards and samples were injected in triplicate. Analytes detected at concentrations below their respective LOQs are reported as detected but not quantified.

Specificity and selectivity were evaluated by comparing chromatograms from methanol blanks, matrix blanks, unspiked product samples, and spiked samples. Methanol served as the solvent blank, and products 5479PR and 5480PR served as representative matrix samples. Spiked samples were prepared by adding known amounts of reference standards to methanol and to the product matrices. Chromatograms of the blank and unspiked samples were examined for interfering peaks at the retention times of the target analytes. Comparison of the chromatographic profiles of the methanol blanks, product samples, and spiked samples showed no significant interfering or co-eluting peaks at the retention times of the analytes. Addition of the reference standards produced well-resolved peaks at the expected retention times, confirming the specificity and selectivity of the method.

Accuracy was evaluated by recovery experiments using two representative products (5479PR and 5480PR) spiked with known amounts of the four reference standards. Samples were evaluated in duplicate at three concentration levels (1, 10, and 20 µg/mL). Percentage recoveries ranged from 97 to 102%, demonstrating acceptable accuracy.

Precision was evaluated for intra-day and inter-day using products 5479PR and 5480PR. Analyses were performed on three consecutive days, with three independent analytical runs per day and duplicate injections for each run. The intra- and inter-day relative standard deviations (RSDs) were below 5%, with a maximum RSD of 2.5%.

The stability of the standard and sample solutions was evaluated at room temperature over 48 hours. Solutions were analyzed initially and at predetermined intervals up to 48 hours. No significant changes in analyte concentrations were observed, indicating that the solutions were stable under the conditions tested.

**References**


[1] Takayama H, Ishikawa H, Kurihara M, et al. Studies on the Synthesis and Opioid Agonistic Activities of Mitragynine-Related Indole Alkaloids: Discovery of Opioid Agonists Structurally Different from Other Opioid Ligands. *J Med Chem.* 2002; 45(9): 1949-1956. doi: 10.1021/jm010576e

[2] Gour A, Mukhopadhyay S, Henderson A, et al. From Kratom to Semi-Synthetic Opioids: The Rise and Risks of MGM-15. *Drug Test Anal.* 2025; 17(12):2384-2389. doi: 10.1002/dta.3952

[3] Avula B, Ali SK, Tatapudi KK, et al. Quantitative analysis of 7-hydroxymitragynine in commercial kratom products and its stability under chemical and physiological conditions. *Phytochem.* 2026; 247:114871. doi: 10.1016/j.phytochem.2026.114871

[4] Stolz A, Navarro V, Hayashi PH, et al. Severe and protracted cholestasis in 44 young men taking bodybuilding supplements: assessment of genetic, clinical and chemical risk factors. *Aliment Pharmacol Ther.* 2019; 49(9): 1195-1204. doi: 10.1111/apt.15211

[5] Crawford C, Avula B, Lindsey AT, et al. Label accuracy of weight loss dietary supplements marketed online with military discounts. *JAMA Network Open*, 2024; *7*(5),e249131. doi: 10.1001/jamanetworkopen.2024.9131

[6] ICH. Validation of Analytical Procedures: Text and Methodology. ICH Harmonized Tripartite Guidelines Nov. 2005

[7] Yuan D, Ma B, Wu C, et al. Alkaloids from the leaves of *Uncaria rhynchophylla* and their inhibitory activity on NO production in lipopolysaccharide-activated microglia. *J Nat Prod*. 2008; 71(7):1271-1274. doi: 10.1021/np8000305

[8] Wang K, Zhou X-Y, Wang Y-Y, et al. Macrophyllionium and Macrophyllines A and B, Oxindole Alkaloids from *Uncaria macrophylla*. *J. Nat Prod*. 2011; 74(1): 12-15. doi: 10.1021/np1004938

[9] Castro SG, Hosoya E, Kitajima M, et al. Uncamarins A–D, oxindole alkaloids from Philippine *Uncaria longiflora* and their anti-amyloidogenic properties. *Phytochem*. 2025; 238: 114585. doi: 10.1016/j.phytochem.2025.114585

[10] Flores-Bocanegra L, Raja HA, Graf TN, et al. The Chemistry of Kratom [*Mitragyna speciosa*]: Updated Characterization Data and Methods to Elucidate Indole and Oxindole Alkaloids. *J Nat Prod.* 2020;83(7):2165-2177. doi: 10.1021/acs.jnatprod.0c00257

**Table S1. Product Identification: Names, Lot or Batch Numbers, and Other Identifying Information**

| #NCNPR | Name | Ingredients per Serving | Source | Batch # (OR) Identifier # | Mfg. Date | Exp. Date | Barcode # |
|---|---|---|---|---|---|---|---|
| 5443PR | SEDA Botanicals Cat's claw_Watermelon squeeze | ***Uncaria tomentosa* - 31.25 mg** | Distributed by: Seds, Tuscon, AZ 85705 | Lot # 6DEE45 | NA | Feb-2026 | 810195102010 |
| 5445PR | Press'd_Strawberry | Per serving: Ascorbic acid-30 mg; Pyridoxine HCl-5mg; Proprietary blend of URIQ COMPLEX **(*Uncaria tomentosa* extract)- 50 mg**; Quercetin-100 mg; *L*-Theanine- 75 mg; Urolithin A - 100 mg | Distributed by: Alternative Ventures, LLC, 1309 Coffeen Avenue, Sheridan, Wyoming 82801 | PBSI-017-26 | NA | NA | 699838604018 |
| 5469PR | Ice Tabz_Strawberry Slush | Ascorbic acid (30 mg); Pyridoxine HCl (5 mg); ***Uncaria tomentosa* extract (37.5 mg);** Quercetin (100 mg); *L*-Theanine (75 mg); Urolithin A (100 mg); | Distributed by: IT Enterprises LLC, 1347 Johnson Lane, Billings, MT 59101 | U2IT14012026SS | Dec-2025 | Dec-2027 | 028395176414 |
| 5470PR | Ice Tabz_Berry Bomb | Ascorbic acid (30 mg); Pyridoxine HCl (5 mg); ***Uncaria tomentosa* extract (37.5 mg);** Quercetin (100 mg); *L*-Theanine (75 mg); Urolithin A (100 mg); | Distributed by: IT Enterprises LLC, 1347 Johnson Lane, Billings, MT 59101 | U2IT14012026BB | Dec-2025 | Dec-2027 | 028395176445 |
| 5471PR | Happy Daze_Watermelon | Ascorbic acid (30 mg); Pyridoxine HCl (5 mg); ***Uncaria tomentosa* extract (12.5 mg);** Quercetin (100 mg); *L*-Theanine (75 mg); Urolithin A (100 mg); | Billings, Montana 59101 | H2PUT22032026W | Apr-2026 | Apr-2028 | 417836521591 |

| | | | | | | | |
|---|---|---|---|---|---|---|---|
| 5472PR | Happy Daze_Blue Razz | Ascorbic acid (30 mg); Pyridoxine HCl (5 mg); ***Uncaria tomentosa* extract (12.5 mg);** Quercetin (100 mg); *L*-Theanine (75 mg); Urolithin A (100 mg); | Billings, Montana 59101 | 2PUT14012026BR | Apr-2026 | Apr-2028 | 417836521577 |
| 5473PR | 7 Max_Klaz_Tropical fruit | ***Uncaria tomentosa* extract (50 mg)** | Produced By: Trinity Labz, Cheyenne, WY 82001. www. metatabz.com | Not available | NA | NA | 602772512233 |
| 5474PR | 7 Max_Klaz_Blue Razz | ***Uncaria tomentosa* extract (50 mg)** | Produced By: Trinity Labz, Cheyenne, WY 82001. www. metatabz.com | Not available | NA | NA | 683723957143 |
| 5475PR | Buzzers_Pink Lemonade | **Cat's claw extract** | Distributed by: Buzzers Co LLC, 1712 Pioneer Ave # 101, Cheyenne, WY 82001 | Not available | NA | NA | 850070153485 |
| 5476PR | Buzzin_Homiez_Grape Crush | **Cat's claw extract** | Distributed by: Buzzers Co LLC, 1712 Pioneer Ave # 101, Cheyenne, WY 82001 | 012626-2-001 | NA | Jan-2028 | 850078706102 |
| 5477PR | Homiez_Rock Sugar | Proprietary Botanical Blend (25 mg); **Other ingredients: Cat's claw extract** | Manufactured By**:** Elevated Enterprises | 041026-1-001 | NA | Apr-2028 | 850050441625 |

| | | | | | | | |
|---|---|---|---|---|---|---|---|
| 5478PR | Homiez_Lemon Drop | Proprietary Botanical Blend (25 mg); **Other ingredients: Cat's claw extract** | Manufactured By: Elevated Enterprises | 041026-1-001 | NA | Apr-2028 | 850050441649 |
| 5479PR | BLNDZ_Grape | ***Uncaria tomentosa* extract (50 mg)** | Manufactured By: Elevated Goods LLC, 1603 Capital Ave, Ste # 426843, Cheyenne, WY 82001 | Not available | NA | NA | 810190664391 |
| 5480PR | Script_Sweet Citrus_Paradise Blend | ***Uncaria tomentosa* extract (Paradise blend) (50 mg)** | Manufactured & Distributed by: Script, 1525 W Orange Grove Ave, Orange, CA 92868 | 460225-8CESC-SC1 | NA | NA | 860015603432 |
| 5481PR | Script_Miami_Labs_Berry Punch_Paradise Blend | ***Uncaria tomentosa* extract (Paradise blend) (50 mg)** | Manufactured & Distributed by: Script, 1525 W Orange Grove Ave, Orange, CA 92868 | 460225-8CESC-BP1 | NA | NA | 860015603401 |
| 5482PR | Script_Peach Sherbet_Paradise Blend | ***Uncaria tomentosa* extract (Paradise blend) (50 mg)** | Manufactured & Distributed by: Script, 1525 W Orange Grove Ave, Orange, CA 92868 | 460225-8CESC-PS1 | NA | NA | 860015603425 |

| | | | | | | | |
|---|---|---|---|---|---|---|---|
| 5483PR | Script_Sour Apple_Paradise Blend | ***Uncaria tomentosa* extract (Paradise blend) (50 mg)** | Manufactured & Distributed by: Script, 1525 W Orange Grove Ave, Orange, CA 92868 | 460225-8CESC-SA1 | NA | NA | 860015603418 |
| 5487PR | Ice Tabz_Blue Razz | Ascorbic acid (30 mg); Pyridoxine HCl (5 mg); ***Uncaria tomentosa* extract (37.5 mg);** Quercetin (100 mg); *L*-Theanine (75 mg); Urolithin A (100 mg); | Distributed by: IT Enterprises LLC, 1347 Johnson Lane, Billings, MT 59101 | U2IT14012026RB | Dec-2025 | Dec-2027 | 028395176438 |
| 5488PR | Ice Tabz_Watermelon Blast | Ascorbic acid (30 mg); Pyridoxine HCl (5 mg); ***Uncaria tomentosa* extract (37.5 mg);** Quercetin (100 mg); *L*-Theanine (75 mg); Urolithin A (100 mg); | Distributed by: IT Enterprises LLC, 1347 Johnson Lane, Billings, MT 59101 | U2IT14012026WB | Dec-2025 | Dec-2027 | 028395176407 |
| 5489PR | Ice Tabz_Cotton Candy | Ascorbic acid (30 mg); Pyridoxine HCl (5 mg); ***Uncaria tomentosa* extract (37.5 mg);** Quercetin (100 mg); *L*-Theanine (75 mg); Urolithin A (100 mg) | Distributed by: IT Enterprises LLC, 1347 Johnson Lane, Billings, MT 59101 | U2IT14012026CC | Dec-2025 | Dec-2027 | 028395176421 |
| 5490PR | Happy Daze_Mistery | Ascorbic acid (30 mg); Pyridoxine HCl (5 mg); ***Uncaria tomentosa* extract (37.5 mg);** Quercetin (100 mg); *L*-Theanine (75 mg); Urolithin A (100 mg); | Billings, Montana 59101 | H2PUT22032026MT | Apr-2026 | Apr-2028 | 417836521614 |

| | | | | | | | |
|---|---|---|---|---|---|---|---|
| 5491PR | Happy Daze_Strawberry | Ascorbic acid (30 mg); Pyridoxine HCl (5 mg); ***Uncaria tomentosa*** **extract (37.5 mg);** Quercetin (100 mg); *L*-Theanine (75 mg); Urolithin A (100 mg); | Billings, Montana 59101 | H2PUT22032026S | Apr-2026 | Apr-2028 | 417836521584 |
| 5492PR | Happy Daze_Mango | Ascorbic acid (30 mg); Pyridoxine HCl (5 mg); ***Uncaria tomentosa*** **extract (37.5 mg);** Quercetin (100 mg); *L*-Theanine (75 mg); Urolithin A (100 mg); | Billings, Montana 59101 | H2PUT22032026M | Apr-2026 | Apr-2028 | 417836521607 |
| 5493PR | Xuno_Mega_Blueberry | **Cat's claw extract,** Kava extract (70 mg) | Distributed by: Kyan Distribution, Lithia Spring, GA | Not available | NA | NA | 676327253735 |
| 5494PR | Tigers Blood_Strawberry_Breeze | ***Uncaria tomentosa*** **(Cat's Claw)** | Manufactured by: ZH Solutions, LLC | TB2CT0727 | NA | NA | 850085992567 |
| 5495PR | Unlimited_Blue Razz | ***Uncaria tomentosa*** **(75 mg)** | Distributed by: Unlimited Tabs, 16056 Heron Ave, La Mirada, CA 90638 | 1103678636004 | NA | Mar-2029 | 110367863641 |
| 5496PR | Unlimited_Cotton Candy | ***Uncaria tomentosa*** **(75 mg)** | Distributed by: Unlimited Tabs, 16056 Heron Ave, La Mirada, CA 90638 | 1103678636005 | NA | Mar-2029 | 110367863658 |

| | | | | | | | |
|---|---|---|---|---|---|---|---|
| 5497PR | Unlimited_Strawberry | ***Uncaria tomentosa* (75 mg)** | Distributed by: Unlimited Tabs, 16056 Heron Ave, La Mirada, CA 90638 | 1103678636006 | NA | Mar-2029 | 110367863665 |
| 5498PR | Unlimited_Watermelon | ***Uncaria tomentosa* (75 mg)** | Distributed by: Unlimited Tabs, 16056 Heron Ave, La Mirada, CA 90638 | 1103678636003 | NA | Mar-2029 | 110367863634 |
| 5499PR | High Heads_Strawberry_Kiwi | Ascorbic acid (30 mg); Pyridoxine HCl (5 mg); ***Uncaria tomentosa* extract (37.5 mg);** Quercetin (100 mg); *L*-Theanine (75 mg); Urolithin A (100 mg); | High Heads Inc., 3616 N Topeka St, Wichita, KS 67219 | HHE100727RR | Jul-2026 | Sep-2027 | 198819566111 |
| 5500PR | High Heads_Watermelon_Pineapple | Ascorbic acid (30 mg); Pyridoxine HCl (5 mg); ***Uncaria* tomentosa extract (37.5 mg);** Quercetin (100 mg); *L*-Theanine (75 mg); Urolithin A (100 mg); | High Heads Inc., 3616 N Topeka St, Wichita, KS 67219 | HHE100727RR | Jul-2026 | Sep-2027 | 744922386810 |
| 5501PR | High Heads_Blueberry_Raspberry | Ascorbic acid (30 mg); Pyridoxine HCl (5 mg); ***Uncaria tomentosa* extract (37.5 mg);** Quercetin (100 mg); *L*-Theanine (75 mg); Urolithin A (100 mg); | High Heads Inc., 3616 N Topeka St, Wichita, KS 67219 | HHE100727RR | Jul-2026 | Sep-2027 | 998799578735 |

| | | | | | | | |
|---|---|---|---|---|---|---|---|
| 5502PR | 7 Max_Klaz_Strawmelon | ***Uncaria tomentosa* extract (50 mg)** | Produced By: Trinity Labz, Cheyenne, WY 82001. www. metatabz.com | Not available | NA | NA | 683723957150 |
| 5503PR | 7 Max_Klaz Very Cherry | ***Uncaria tomentosa* extract (50 mg)** | Produced By: Trinity Labz, Cheyenne, WY 82001. www. metatabz.com | Not available | NA | NA | 602772512240 |
| 5504PR | 7 Max_Klaz_Great Grape | ***Uncaria tomentosa* extract (50 mg)** | Produced By: Trinity Labz, Cheyenne, WY 82001. www. metatabz.com | Not available | NA | NA | 683723957167 |
| 5505PR | Blend_Watermelon | ***Uncaria tomentosa* (50 mg)** | Manufactured & Distributed by: Blend Products, 10005 Muirlands Blvd., Irvine, CA 92618 | Not available | NA | NA | 850083811662 |
| 5506PR | Blend_Strawberry | ***Uncaria tomentosa* (50 mg)** | Manufactured & Distributed by: Blend Products, 10005 Muirlands Blvd., Irvine, CA 92618 | Not available | NA | NA | 850083811655 |
| 5507PR | Blend_Blue Razz | ***Uncaria tomentosa* (50 mg)** | Manufactured & Distributed by: Blend Products, | Not available | NA | NA | 850083811631 |

| | | | | | | | |
|---|---|---|---|---|---|---|---|
| | | | 10005 Muirlands Blvd., Irvine, CA 92618 | | | | |
| 5508PR | Script_Sweet Citrus_Paradise Blend | ***Uncaria tomentosa* extract (50 mg)** | Manufactured & Distributed by: Script, 1525 W Orange Grove Ave, Unit B, Orange, CA 92868 | Not available | NA | NA | 860015329431 |
| 5509PR | 7 Synergy_Fruit Punch | ***Uncaria tomentosa* extract (50 mg)** | Distributed by: 7 Synergy Distribution, 16056 Heron Ave, La Mirada, CA 90638 | 110463786228 | NA | Mar-2029 | 123456245112 |
| 5510PR | 7 Synergy_Blue Razz | ***Uncaria tomentosa* extract (50 mg)** | Distributed by: 7 Synergy Distribution, 16056 Heron Ave, La Mirada, CA 90638 | 110463786232 | NA | Mar-2029 | 123456245518 |
| 5511PR | 7 Synergy_Watermelon Drop | ***Uncaria tomentosa* extract (50 mg)** | Distributed by: 7 Synergy Distribution, 16056 Heron Ave, La Mirada, CA 90638 | 110463786230 | NA | Mar-2029 | 123456245310 |
| 5512PR | Drift_Blue Razz | ***Uncaria tomentosa* extract (250 mg)** | Distributed by: Drift | drift-012 | NA | Jul-2028 | 732262073095 |

| | | | | | | | |
|---|---|---|---|---|---|---|---|
| | | | Botanicals, Miami, FL 33130 | | | | |
| 5513PR | Drift_Berry Flavour | ***Uncaria tomentosa* extract (250 mg)** | Distributed by: Drift Botanicals, Miami, FL 33130 | drift-012 | NA | Jul-2028 | 732262073088 |
| 5514PR | Drift_Pink Strawberry | ***Uncaria tomentosa* extract (250 mg)** | Distributed by: Drift Botanicals, Miami, FL 33130 | drift-012 | NA | Jul-2028 | 732262073118 |
| 5515PR | Drift_Purple Grape | ***Uncaria tomentosa* extract (250 mg)** | Distributed by: Drift Botanicals, Miami, FL 33130 | drift-012 | NA | Jul-2028 | 732262073125 |

**Table S2. Quantitative Analysis of Target Alkaloids and Compounds Across Sample Products** (mg/unit dosage form)

| # | NCNPR Code # | Mitragynine | 7-Hydroxy mitragynine | Mitragynine pseudoindoxyl | 3-Dehydro mitragynine | MGM-15 | Tianeptine | Caffeine |
|---|---|---|---|---|---|---|---|---|
| 1 | 5443PR | 0.02±2.1 | ND | ND | <LOQ | ND | ND | ND |
| 2 | 5445PR | 0.8±0.9 | ND | ND | 0.03±2.1 | ND | ND | ND |
| 3 | 5469PR | 0.4±0.3 | 73.2±0.1 | 0.6±0.6 | 0.4±0.9 | ND | ND | ND |
| 4 | 5470PR | 0.4±0.7 | 45.2±0.1 | 0.4±0.7 | 1.01±0.3 | ND | ND | ND |
| 5 | 5471PR | 5.6±0.1 | 45.3±0.2 | 0.5±0.6 | 1.7±0.3 | ND | ND | ND |
| 6 | 5472PR | ND | 0.01±0.7 | ND | 0.01±1.1 | ND | 27.9 ± 0.06 | 0.11±1.3 |
| 7 | 5473PR | 1.3±0.3 | 0.01±1.9 | 0.1±2.2 | 0.1±0.9 | 22.23±0.6 | ND | ND |
| 8 | 5474PR | 1.6±0.1 | 0.01±1.4 | 0.1±1.4 | 0.1±0.9 | 8.12±0.1 | ND | 0.14±1.5 |
| 9 | 5475PR | 0.4±0.7 | ND | ND | 0.1±2.0 | 4.72±0.2 | ND | ND |
| 10 | 5476PR | 1.1±0.3 | 0.5±0.4 | 0.1±0.3 | 0.2±1.4 | 7.5±0.03 | ND | ND |
| 11 | 5477PR | 0.6±0.6 | 0.04±1.8 | ND | 0.05±0.3 | 6.22±0.1 | ND | ND |
| 12 | 5478PR | 0.6±0.7 | 0.1±0.1 | ND | 0.06±0.2 | 5.7±0.1 | ND | ND |
| 13 | 5479PR | 1.1±0.2 | 0.04±0.4 | ND | 0.03±0.4 | 16.3±0.03 | ND | ND |
| 14 | 5480PR | 0.8±0.4 | ND | ND | 0.3±0.8 | 23.0±0.01 | ND | 3.2±0.3 |
| 15 | 5481PR | 0.9±0.3 | ND | ND | 0.1±2.5 | 25.7±0.01 | ND | 3.1±0.5 |
| 16 | 5482PR | 0.9±0.8 | ND | ND | 0.04±0.9 | 25.6±0.2 | ND | 3.3±0.1 |
| 17 | 5483PR | 1.1±0.1 | ND | ND | 0.1±2.6 | 25.4±0.2 | ND | 3.2±0.2 |
| 18 | 5487PR | 0.1±1.2 | 84.2±0.2 | 0.8±0.4 | 0.4±0.9 | ND | ND | ND |
| 19 | 5488PR | 0.4±0.9 | 53.2±0.2 | 0.5±0.6 | 0.3±0.5 | ND | ND | ND |
| 20 | 5489PR | 0.1±1.3 | 78.4±0.1 | 0.7±0.4 | 0.3±0.9 | ND | ND | ND |
| 21 | 5490PR | 0.6±0.6 | 75.4±0.1 | 0.6±0.5 | 2.03±0.3 | ND | ND | ND |
| 22 | 5491PR | 0.4±0.9 | 45.8±0.02 | 0.4±1 | 1.02±0.6 | ND | ND | ND |
| 23 | 5492PR | ND | 0.01±0.6 | ND | 0.01±1.9 | ND | 28.2±0.2 | 0.1±0.9 |
| 24 | 5493PR | 0.1±0.1 | 0.03±2.2 | ND | 0.02±3.1 | 13.2±0.5 | ND | 0.5±0.6 |

| | | | | | | | | |
|---|---|---|---|---|---|---|---|---|
| 25 | 5494PR | 1.4±0.3 | 0.01±1.5 | ND | 0.1±2.5 | 23.2±0.1 | ND | ND |
| 26 | 5495PR | 0.8±0.5 | 22.2±0.3 | 0.7±0.3 | 0.6±0.5 | ND | ND | ND |
| 27 | 5496PR | 0.4±0.9 | 37.5±0.2 | 1.2±0.1 | 1.4±0.3 | ND | ND | ND |
| 28 | 5497PR | 0.4±0.9 | 40.7±0.1 | 0.8±0.4 | 1.4±0.2 | ND | ND | ND |
| 29 | 5498PR | 0.5±0.3 | 40.8±0.01 | 1.00±0.6 | 1.3±0.3 | ND | ND | ND |
| 30 | 5499PR | ND | 0.1±0.9 | ND | 0.01±0.6 | ND | 40.2±0.4 | 0.1±0.9 |
| 31 | 5500PR | ND | 0.1±1.4 | ND | 0.02±0.5 | ND | 39.3±0.4 | 0.1±1.9 |
| 32 | 5501PR | ND | 0.02±1.3 | ND | 0.01±1.3 | ND | 39.5±0.9 | ND |
| 33 | 5502PR | 0.5±0.1 | 0.01±0.1 | ND | 0.02±2.7 | 21.0±0.6 | ND | 3.5±0.2 |
| 34 | 5503PR | 0.1±1.2 | 0.04±4.9 | ND | 0.01±1.3 | 28.4±0.4 | ND | 2.5±0.3 |
| 35 | 5504PR | 0.1±0.6 | 0.08±0.9 | ND | 0.1±1.2 | 20.3±1.2 | ND | 3.4±0.9 |
| 36 | 5505PR | 2.6±0.1 | ND | ND | 0.2±1.1 | 18.6±0.4 | ND | ND |
| 37 | 5506PR | 1.3±0.3 | ND | ND | 0.3±0.7 | 27.9±0.2 | ND | ND |
| 38 | 5507PR | 1.3±0.1 | ND | ND | 0.07±1 | 14.5±0.5 | ND | ND |
| 39 | 5508PR | 1.5±2.1 | ND | ND | 0.01±2.8 | 27.6±0.4 | ND | 2.7±1.4 |
| 40 | 5509PR | 0.4±0.7 | 37.6±0.1 | 1.5±0.1 | 1.5±0.5 | ND | ND | ND |
| 41 | 5510PR | 32.5±0.2 | 0.1±0.5 | 0.03±0.7 | 0.3±0.9 | 27.8±0.2 | ND | ND |
| 42 | 5511PR | 0.3±0.2 | 38.4±0.7 | 0.96±0.3 | 1.3±0.5 | ND | ND | ND |
| 43 | 5512PR | 1.4±0.1 | 95.4±0.2 | 1.1±0.2 | 1.8±0.1 | ND | ND | ND |
| 44 | 5513PR | 1.6±0.3 | 79.5±0.01 | 1.5±0.2 | 2.5±0.1 | 3.1±0.1 | ND | ND |
| 45 | 5514PR | 31.2±0.1 | 0.7±0.4 | 1.4±0.3 | 0.5±0.7 | 24.8±0.2 | ND | ND |
| 46 | 5515PR | 31.7±0.1 | 0.8±0.2 | 1.7±0.1 | 0.4±0.3 | 28.7±0.3 | ND | ND |

**Note:** Values are presented as mean ± RSD (n = 3). ND, not detected.

**Figure S1. Composition of measured compounds in each of 46 products labeled as cat's claw.** Each pie shows the product's summed mg per unit appears under each pie.

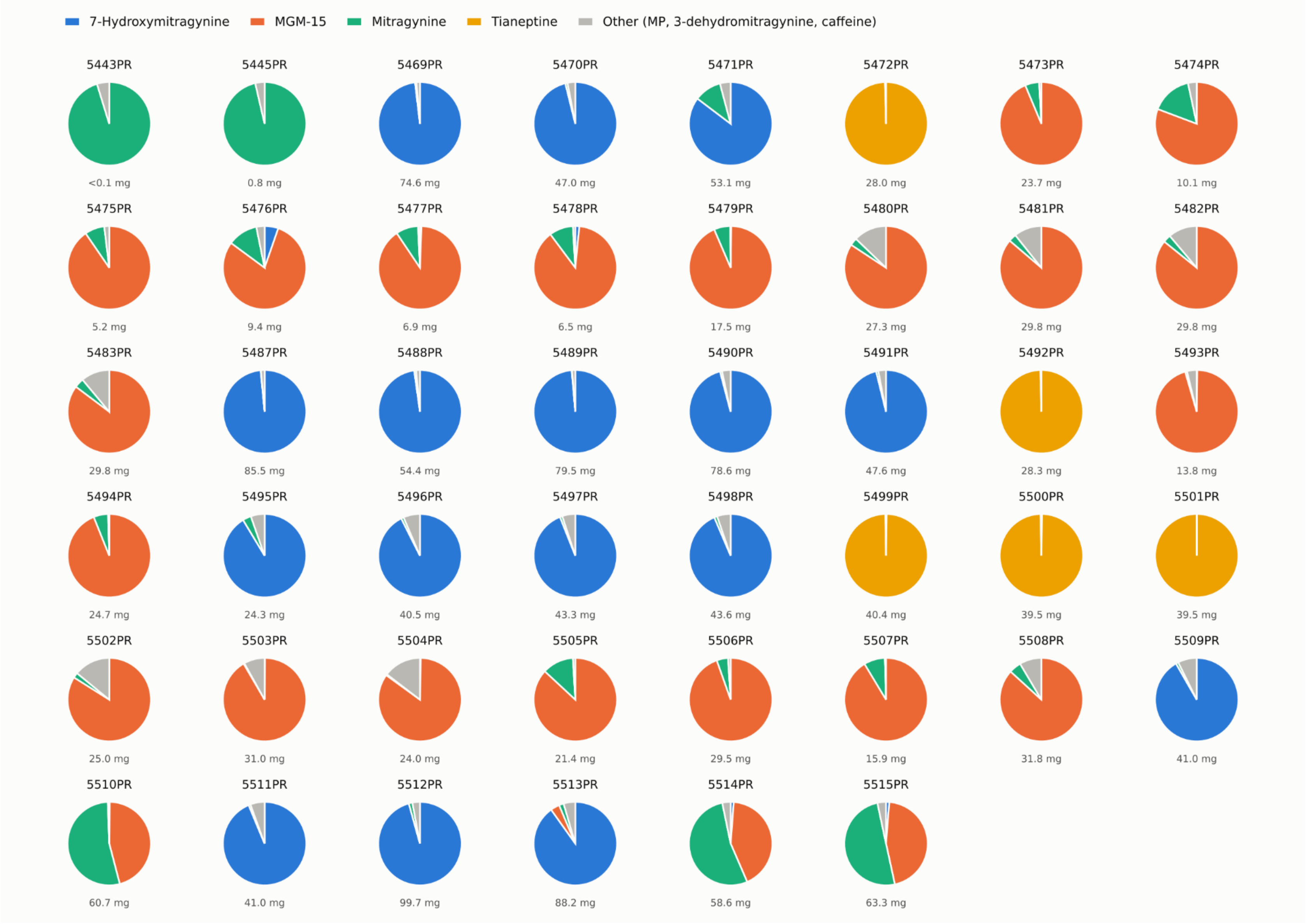

**Table S3. LC–QToF–MS identification and characterization of compounds in commercial products marketed as cat's claw (*Uncaria tomentosa*).**

| # | Retention time ($t_R$, min) | Compound | Molecular formula | Exact mass | *m/z* $[M+H]^+$ | Key fragment ions (*m/z*)† | Product code(s) |
|---|---|---|---|---|---|---|---|
| ***Oxindole Alkaloids of Uncaria tomentosa*** | | | | | | | |
| 1 | 18.26 | Uncarine E (= Isopteropodine) | $C_{21}H_{24}N_2O_4$ | 368.1736 | 369.1811 (369.1809) | 337.1549, 201.1014, **160.0757** | 5476PR; 5478PR; 5493PR; 5494PR; 5506PR |
| 2 | 18.73 | Isomitraphylline | $C_{21}H_{24}N_2O_4$ | 368.1736 | 369.1813 (369.1809) 391.1633 (391.1628) $[M+Na]^+$ | 337.1554, 309.1603, 178.0858, **160.0757** | |
| 3 | 18.92 | Mitraphylline | $C_{21}H_{24}N_2O_4$ | 368.1736 | 369.1814 (369.1809) 391.1627 (391.1628) $[M+Na]^+$ | 337.1548, 309.1601, 178.0857, **160.0757** | |
| 4 | 19.39 | Uncarine D (= Speciophylline) | $C_{21}H_{24}N_2O_4$ | 368.1736 | 369.1814 (369.1809) 391.1626 (391.1628) $[M+Na]^+$ | 337.1552, 201.1021, **160.0750**, 158.0588 | |
| 5 | 21.23 | Uncarine C (= Pteropodine) | $C_{21}H_{24}N_2O_4$ | 368.1736 | 369.1805 (369.1809) | 337.1550, 201.1024, **160.0750** | |
| **Corynanthe-type indole alkaloids (*Mitragyna*/kratom-related)** | | | | | | | |
| 6 | 23.98 | Corynantheidine | $C_{22}H_{28}N_2O_3$ | 368.2100 | 369.2175 (369.2173) | 337.1916 $[M+H-CH_4O]^+$, 238.1445 $[C_{13}H_{20}NO_3]^+$, 226.1442 $[C_{12}H_{20}NO_3]^+$, **144.0806** $[C_{10}H_{10}N]^+$ | 5469PR; 5470PR; 5471PR; 5473PR; 5474PR; 5475PR; 5476PR; 5477PR; 5478PR; 5479PR; 5480PR; 5481PR; 5482PR; 5483PR; 5491PR, 5493PR-5497PR; 5500PR, 5502PR; 5505PR-5509PR; 5511PR-5513PR |
| 7 | 25.12 | Paynantheine | $C_{23}H_{28}N_2O_4$ | 396.2049 | 397.2125 (397.2122) | 238.1425 $[C_{13}H_{20}NO_3]^+$, **224.1284** $[C_{12}H_{18}NO_3]^+$, **174.0913** $[C_{11}H_{12}NO]^+$ | 5471PR; 5473PR; 5474PR; 5475PR; 5476PR; 5477PR; 5478PR; 5479PR; 5480PR-5483PR; 5491PR; 5493PR-5498PR; 5502PR; 5505PR-5515PR |

| # | Retention time ($t_R$, min) | Compound | Molecular formula | Exact mass | *m/z* [M+H]⁺ | Key fragment ions (*m/z*)† | Product code(s) |
|---|---|---|---|---|---|---|---|
| 8 | 25.23 | Mitragynine | $C_{23}H_{30}N_2O_4$ | 398.2206 | 399.2282 (399.2278) | 367.2021 $[M+H-CH_4O]^+$, 238.1435 $[C_{13}H_{20}NO_3]^+$, **226.1432** $[C_{12}H_{20}NO_3]^+$, **174.0912** $[C_{11}H_{12}NO]^+$ | 5443PR; 5445PR; 5469PR; 5470PR; 5471PR; 5473PR; 5474PR; 5475PR; 5476PR; 5477PR; 5478PR; 5479PR; 5480PR; 5481PR; 5482PR; 5483PR; 5487PR; 5488PR; 5489PR; 5490PR; 5491PR; 5493PR-5498PR; 5502PR-5515PR |
| 9 | 25.99 | Speciogynine | $C_{23}H_{30}N_2O_4$ | 398.2206 | 399.2277 (399.2278) | 367.2026 $[M+H-CH_4O]^+$, 238.1441 $[C_{13}H_{20}NO_3]^+$, **226.1438** $[C_{12}H_{20}NO_3]^+$, **174.0911** $[C_{11}H_{12}NO]^+$ | 5471PR; 5473PR; 5474PR; 5475PR; 5476PR; 5477PR; 5478PR; 5479PR; 5480PR-5483PR; 5491PR; 5493PR-5498PR; 5502PR; 5505PR-5515PR |
| 10 | 26.10 | Corynantheine | $C_{22}H_{26}N_2O_3$ | 366.1943 | 367.2019 (367.2016) | **224.1281** $[C_{12}H_{18}NO_3]^+$, **144.0800** $[C_{10}H_{10}N]^+$ | 5469PR; 5470PR; 5471PR; 5473PR; 5474PR; 5445PR |
| 11 | 26.76 | Speciociliatine | $C_{23}H_{30}N_2O_4$ | 398.2206 | 399.2281 (399.2278) | 367.2017 $[M+H-CH_4O]^+$, 238.1420 $[C_{13}H_{20}NO_3]^+$, 226.1434 $[C_{12}H_{20}NO_3]^+$, **174.0910** $[C_{11}H_{12}NO]^+$ | 5471PR; 5473PR; 5474PR; 5475PR; 5476PR; 5477PR; 5478PR; 5479PR; 5480PR-5483PR; 5491PR; 5493PR-5498PR; 5502PR; 5505PR-5515PR |
| **Mitragynine-related oxygenated, oxidized, and semisynthetic derivatives** | | | | | | | |
| 12 | 18.65 | 7-Hydroxymitragynine | $C_{23}H_{30}N_2O_5$ | 414.2155 | 415.2230 (415.2227) | 397.2133 $[M+H-H_2O]^+$, 238.1431 $[C_{13}H_{20}NO_3]^+$, **190.0867** $[C_{11}H_{12}NO_2]^+$ | 5469PR; 5470PR; 5471PR; 5472PR; 5473PR; 5474PR; 5476PR; 5477PR; 5478PR; 5479PR; 5487PR; 5488PR; 5489PR; 5490PR; 5491PR; 5492PR-5504PR; 5509PR-5515PR |
| 13 | 20.59 | Dihydro-7-hydroxy mitragynine (**MGM-15**) | $C_{23}H_{32}N_2O_5$ | 416.2311 | 417.2381 (417.2384) | **399.2265** $[M+H-H_2O]^+$, **192.1018** $[C_{11}H_{14}NO_2]^+$ | 5473PR; 5474PR; 5475PR; 5476PR; 5477PR; 5478PR; 5479PR; 5480PR; 5481PR; 5482PR; 5483PR; 5493PR; 5494PR;5502PR-5508PR; 5510PR; 5513PR-5515PR |
| 14 | 21.69 | Mitragynine pseudoindoxyl | $C_{23}H_{30}N_2O_5$ | 414.2155 | 415.2225 (415.2227) | **383.1977** $[M+H-CH_4O]^+$, 238.1435 $[C_{13}H_{20}NO_3]^+$, 190.0861 $[C_{11}H_{12}NO_2]^+$, **150.0918** $[C_9H_{12}NO]^+$ | 5469PR; 5470PR; 5471PR; 5473PR; 5474PR; 5476PR; 5487PR; 5488PR; 5489PR; 5490PR; 5491PR; 5495PR-5498PR; 5509PR; 5510PR; 5511PR-5515PR |
| 15 | 25.52 | | $C_{23}H_{30}N_2O_5$ | 414.2155 | 415.2226 | | 5510PR; 5514PR; 5515PR |

| # | Retention time ($t_R$, min) | Compound | Molecular formula | Exact mass | $m/z$ $[M+H]^+$ | Key fragment ions ($m/z$)† | Product code(s) |
|---|---|---|---|---|---|---|---|
| 16 | 27.19 | Mitragynine *N*-oxide isomers* | | | (415.2227) | 399.2276, 397.2127 $[M+H-H_2O]^+$, 383.2114 $[M+H-CH_2OH]^+$, 369.1961, 367.2019, 353.1883, 337.1914, **174.0912** | 5469PR; 5470PR; 5479PR; 5487PR; 5488PR; 5489PR; 5494PR; 5495PR; 5496PR; 5497PR; 5506PR; 5507PR; 5509PR; 5510PR; 5511PR; 5514PR; 5515PR |
| 17 | 28.05 | 3-Dehydromitragynine | $C_{23}H_{28}N_2O_4$ | 396.2049 | 397.2127 (397.2122) | 365.1854 $[M+H-CH_4O]^+$, 236.1273 $[C_{13}H_{18}NO_3]^+$, 224.1270 $[C_{12}H_{18}NO_3]^+$, **172.0753** $[C_{11}H_{10}NO]^+$ | 5443PR; 5445PR; 5469PR–5483PR; 5487PR–5515PR |
| **Other indole derivatives** | | | | | | | |
| 18 | 19.09 | Yohimbine | $C_{21}H_{26}N_2O_3$ | 354.1943 | 355.2019 (355.2016) | 212.1285 $[C_{11}H_{18}NO_3]^+$, **144.0807** $[C_{10}H_{10}N]^+$ | 5443PR |
| **Spirooxindole alkaloids** | | | | | | | |
| 19 | 19.58 | Hydroxy-rhynchophylline isomer* | $C_{22}H_{28}N_2O_5$ | 400.1998 | 401.2075 (401.2071) | 383.1982 $[M+H-H_2O]^+$, 369.1818 $[M+H-CH_4O]^+$, 285.1600 $[M+H-C_5H_8O_3]^+$, 203.0809 $[C_{11}H_{11}N_2O_2]^+$, **176.0701** $[C_{10}H_{10}NO_2]^+$ | 5445PR; 5471PR; 5491PR; |
| 20 | 20.73 | Rhynchophylline# | $C_{22}H_{28}N_2O_4$ | 384.2049 | 385.2123 (385.2122) | 353.1869 $[M+H-CH_4O]^+$, 269.1664 $[M+H-C_5H_8O_3]^+$, 241.1332 $[C_{15}H_{17}N_2O]^+$, **160.0748** $[C_{10}H_{10}NO]^+$ | 5445PR; 5471PR; 5474PR; 5495PR-5498PR; 5502PR; 5507PR-5509PR; 5512PR; 5513PR |
| 21 | 23.71 | Corynoxeine* | $C_{22}H_{26}N_2O_4$ | 382.1893 | 383.1965 (383.1965) | 351.1712 $[M+H-CH_4O]^+$, **267.1482** $[M+H-C_5H_8O_3]^+$, 239.1171 $[C_{15}H_{15}N_2O]^+$, **160.0758** $[C_{10}H_{10}NO]^+$ | 5445PR |
| **Other non-indole compounds and pharmaceutical agents** | | | | | | | |
| 22 | 2.77 | Theanine | $C_7H_{14}N_2O_3$ | 174.1004 | 175.1078 (175.1077) | 158.0813 $[M+H-NH_3]^+$, 130.0498 $[M+H-C_2H_7N]^+$, 102.0541 $[M+H-C_2H_7N-CO]^+$ | 5475PR; 5476PR; 5493PR; 5503PR; 5506PR |

| # | Retention time ($t_R$, min) | Compound | Molecular formula | Exact mass | *m/z* $[M+H]^+$ | Key fragment ions (*m/z*)† | Product code(s) |
|---|---|---|---|---|---|---|---|
| 23 | 13.38 | Caffeine | $C_8H_{10}N_4O_2$ | 194.0804 | 195.0878 (195.0877) | **138.0667** $[M+H-C_2H_3NO]^+$, 123.0441 $[M+H-C_2H_3NO-CH_3]^+$, 110.0716 $[M+H-C_2H_3NO-CO]^+$ | 5472PR; 5474PR; 5480PR; 5481PR; 5482PR; 5483PR; 5492PR; 5493PR; 5499PR; 5500PR, 5502PR-5504PR; 5508PR |
| 24 | 15.05 | Lidocaine | $C_{14}H_{22}N_2O$ | 234.1732 | 235.1801 (235.1805) | 162.0912 $[M+H-C_4H_{11}N]^+$, **86.0956** $[C_5H_{12}N]^+$, 58.0642 $[C_5H_{12}N-C_2H_4]^+$ | 5443PR |
| 25 | 20.13 | Melatonin | $C_{13}H_{16}N_2O_2$ | 232.1212 | 233.1288 (233.1285) | 216.1022 $[M+H-NH_3]^+$, **174.0913** $[M+H-C_2H_5NO]^+$, 159.0680 $[M+H-C_2H_5NO-CH_3]^+$, 130.0646 $[C_9H_8N]^+$ | 5443PR |
| 26 | 24.08 | Tianeptine | $C_{21}H_{25}ClN_2O_4S$ | 436.1224 | 437.1299 (437.1296) | **292.0194** $[C_{14}H_{11}ClNO_2S]^+$, **228.0559** $[C_{14}H_{11}ClN]^+$, 193.0870 $[C_{14}H_{11}N]^+$ | 5472PR; 5492PR; 5499PR-5501PR |

NOTE: *Tentative identification based on its accurate mass and fragmentation; †Bold numbers represent diagnostic major fragment ions; Values in parentheses represent calculated *m/z* values for $[M+H]^+$; Compounds without an asterisk were confirmed by comparison of retention time and MS/MS spectra with authentic reference standards.# Rhynchophylline is not specific to cat's claw, because it has also been reported in *Uncaria spp*[7-9] and *Mitragyna speciosa*[10], so its detection alone cannot identify the source plant.
